# When should healthcare workers with COVID-19 return to work? An analysis of follow-up antigen test results after a positive COVID test

**DOI:** 10.1101/2022.09.24.22280327

**Authors:** Anthony Baffoe-Bonnie, Mandy C. Swann, Hyun Sue Kim

## Abstract

**Background:** Under CDC’s guidance for mitigating healthcare worker (HCW) staffing shortages, COVID-positive HCW may return to work as early as five days after their initial positive test without a negative antigen test, if symptoms are improving. Recent studies suggest a robust correlation between a positive COVID-19 antigen test and infectiousness.

**Methods:** From January to June 2022, HCW employed by a large health system who tested positive for COVID on a PCR test were instructed to isolate and return for a rapid antigen test on day 5 or later if they had been fever-free for 24 hours and their symptoms were improving. We conducted chi-squared tests and a multivariate logistic regression to assess the association between demographic characteristics, vaccination status, and days from the initial positive PCR test on RTW antigen test results.

**Results:** Compared to day 5, HCW had a lower odds of a positive antigen result on day 7 (OR: 0.39, p<0.0001) and after at least 8 days (OR: 0.16, p<0.0001). Unexpectedly, a positive antigen result was more likely among HCW who were vaccinated (OR: 1.41, p <0.05), boosted for more than 90 days prior (OR: 2.21, p<0.0001), and boosted within 90 days (OR: 2.08, p < 0.01) compared to not being vaccinated.

**Conclusions:** Our findings suggest that HCW returning to work before day 7 following a positive PCR test may still be infectious and future guidelines addressing contingency staffing should reflect these findings in order to minimize possible transmission in the healthcare setting. The finding that boosted individuals had over twice the odds of returning positive on the follow up antigen test compared to unvaccinated HCWs merits additional research.

## Background

The Centers for Disease Control and Prevention’s (CDC) guidance for work restriction and the duration of isolation for healthcare workers (HCWs) after a COVID-19 infection has evolved throughout the pandemic ^1^. The current guidance outlines a continuum of options— from conventional to contingency to crisis— that suggest varying durations of work restriction for the immunocompetent HCW who has an asymptomatic infection or where symptoms are improving following a mild or moderate COVID-19 infection ^2^. This guidance aims to minimize HCW staffing shortages while mitigating the risk of SARS-CoV-2 transmission in the healthcare setting. Under the contingency standards, COVID-19 positive HCWs may return to work after five days with or without a negative test ^1,3,4^.

Preliminary findings from a study of a single urban center during the omicron surge noted a high rate of positive antigen results in HCWs who were tested to end isolation on day five following a positive PCR COVID test ^5^. Similarly, the Yukon-Kuskokwim Health Corporation (YKHC) in Alaska noted that amongst persons in the community who had symptomatic COVID-19 and returned for repeat testing on day 5 after symptom onset, the antigen positivity rate was 80 percent ^6^. There is now accruing evidence suggesting a strong correlation between a positive COVID-19 antigen test and positive viral cultures or infectivity ^4,7,8^. The implications of these findings merit continued research in this area to guide return to work recommendations for healthcare workers and the population at large.

We report the HCW return-to-work (RTW) experience of a large health system in southwest Virginia at a time when the predominant strain was the Omicron variant and its subvariants. Our study examines factors associated with a positive RTW antigen test while determining if the findings from prior studies are replicable in this different HCW population and with different subvariants.

## Methods

We conducted a retrospective cross-sectional study of employees of a large healthcare system in southwest Virginia who tested positive for COVID-19 and completed RTW antigen testing between January 6, 2022, and July 6, 2022. Symptomatic employees were instructed to isolate and immediately undergo a SARS-CoV-2 RT-PCR test performed by trained personnel at designated testing centers. The day the positive PCR test was collected was designated as day zero for RTW purposes and guide the RTW timeline.

SARS-CoV-2 PCR-positive employees were asked to continue isolation and return for a single RTW COVID-19 antigen test (Quidel Sofia SARS Antigen FIA) obtained by trained personnel on day 5 or later if they had been fever-free for 24 hours without an antipyretic and their symptoms were improving. If an employee had a positive RTW antigen test, they would not retest but could return after day 10 if they were afebrile for the preceding 24 hours (Appendix A). Employees who were immunocompromised or had severe/critical COVID-19 disease were not eligible to utilize this protocol.

Data on gender, age, vaccine type, date of vaccination, the interval in days between the positive COVID-19 PCR test and the RTW antigen test, and the result of the RTW antigen test were obtained through the electronic administrative records of our centralized contact tracing team. This study was deemed exempt by the Carilion Clinic IRB.

Vaccination status was defined using the number and type of vaccines completed at least 14 days before the initial positive SARS-CoV-2 RT PCR test. HCWs who had not received any COVID-19 vaccine or had only one dose of an mRNA vaccine were considered “not vaccinated”. HCWs who had received either a single dose of the Johnson & Johnson/Janssen (J&J) vaccine or two doses of an mRNA vaccine were considered vaccinated. HCWs who had received two J&J doses, an mRNA vaccine following a J&J shot, or three or more mRNA vaccines were considered boosted. To assess the timing of the booster on the RTW antigen results, we subclassified boosted HCW into 2 categories: those boosted less than 90 days before their initial positive PCR test and those boosted 90 or more days before their PCR test.

All records where the completion of RTW antigen testing was performed between day 5 and day 11 after the initial positive PCR test (Day 0) were analyzed. Categorical data were described as frequencies and percentages and compared by chi-squared tests. A logistic regression model was examined to identify characteristics independently associated with a positive antigen result while controlling for characteristics included in the model. Each characteristic associated with antigen results (p<0.05) was considered an explanatory variable in the model. All analyses were completed using SAS version 9.4 (SAS Institute, Cary, NC).

## Results

Throughout the study period, 1,704 HCWs completed RTW antigen testing following a positive SARS-CoV-2 RT-PCR test. Table 1 outlines the descriptive characteristics of these HCWs and presents the results of chi-squared tests.

**TABLE 1:**
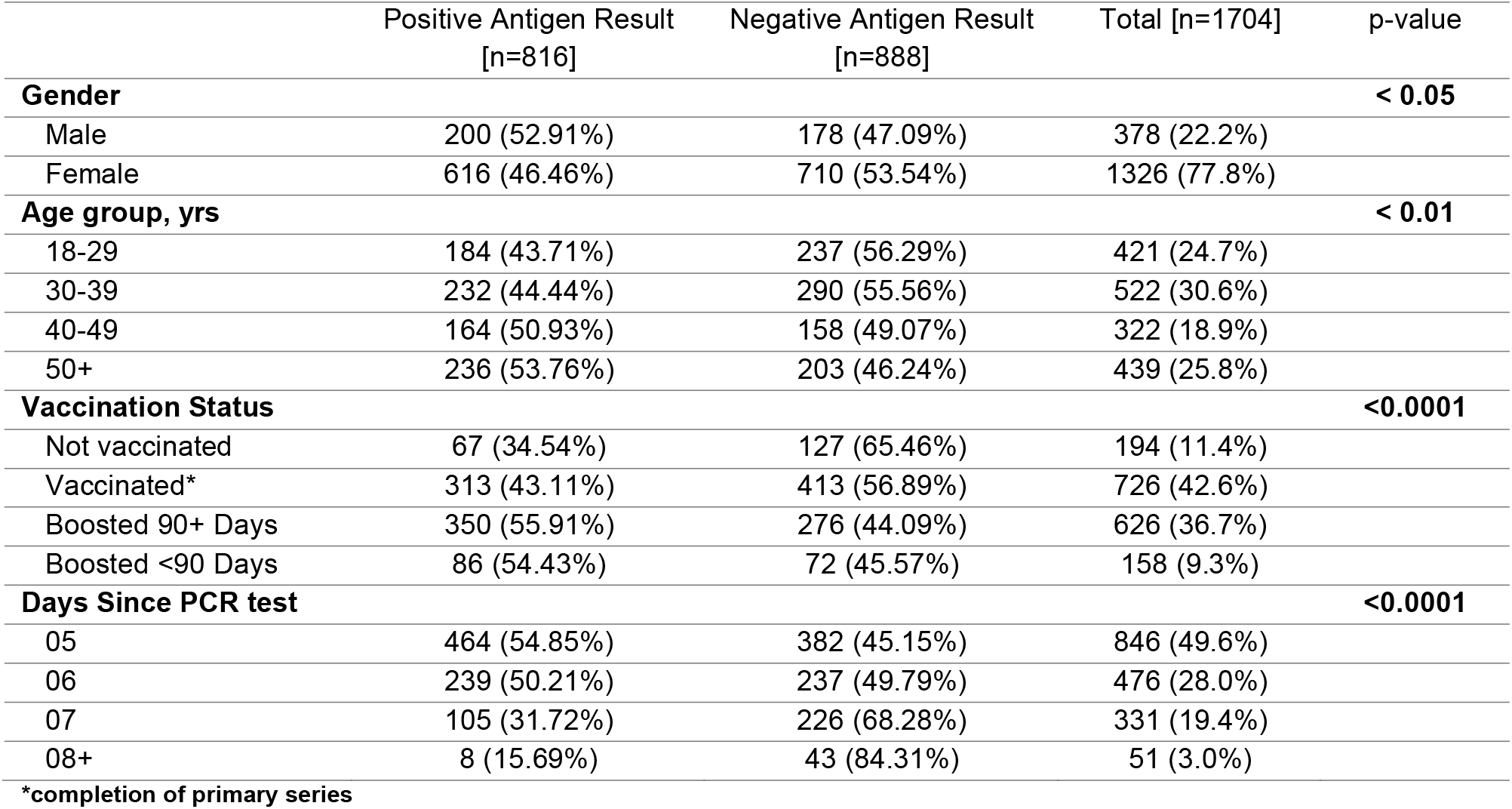
Descriptive characteristics of symptomatic HCWs completing RTW antigen testing following a positive SARS-CoV-2 RT-PCR result.

|  | Positive Antigen Result<br>[n=816] | Negative Antigen Result<br>[n=888] | Total [n=1704] | p-value |
| --- | --- | --- | --- | --- |
| **Gender** |  |  |  | **< 0.05** |
| Male | 200 (52.91%) | 178 (47.09%) | 378 (22.2%) |  |
| Female | 616 (46.46%) | 710 (53.54%) | 1326 (77.8%) |  |
| **Age group, yrs** |  |  |  | **< 0.01** |
| 18-29 | 184 (43.71%) | 237 (56.29%) | 421 (24.7%) |  |
| 30-39 | 232 (44.44%) | 290 (55.56%) | 522 (30.6%) |  |
| 40-49 | 164 (50.93%) | 158 (49.07%) | 322 (18.9%) |  |
| 50+ | 236 (53.76%) | 203 (46.24%) | 439 (25.8%) |  |
| **Vaccination Status** |  |  |  | **<0.0001** |
| Not vaccinated | 67 (34.54%) | 127 (65.46%) | 194 (11.4%) |  |
| Vaccinated* | 313 (43.11%) | 413 (56.89%) | 726 (42.6%) |  |
| Boosted 90+ Days | 350 (55.91%) | 276 (44.09%) | 626 (36.7%) |  |
| Boosted <90 Days | 86 (54.43%) | 72 (45.57%) | 158 (9.3%) |  |
| **Days Since PCR test** |  |  |  | **<0.0001** |
| 05 | 464 (54.85%) | 382 (45.15%) | 846 (49.6%) |  |
| 06 | 239 (50.21%) | 237 (49.79%) | 476 (28.0%) |  |
| 07 | 105 (31.72%) | 226 (68.28%) | 331 (19.4%) |  |
| 08+ | 8 (15.69%) | 43 (84.31%) | 51 (3.0%) |  |

\*completion of primary series

The majority of HCWs in our sample were female (78%) and the mean age was 40 years. 89% of the healthcare workers were either fully vaccinated or boosted at the time of their initial PCR test. Half (50%) of eligible HCW completed RTW testing on day 5 following their initial positive PCR test and 97% had tested by day 7. Nearly half (48%) of the HCWs had a positive result on their return-to-work antigen test. The results of the chi-squared tests indicate that gender, age, vaccination status, and days since initial positive PCR were all significantly associated with the RTW antigen test result (Table 1).

Figure 1 depicts the percent positivity of RTW antigen tests and 95% confidence intervals by the number of days since the initial PCR test. Tests completed before day 7 were significantly more likely to be positive than those done on day 7 or later (p < 0.05).

**Figure 1:**
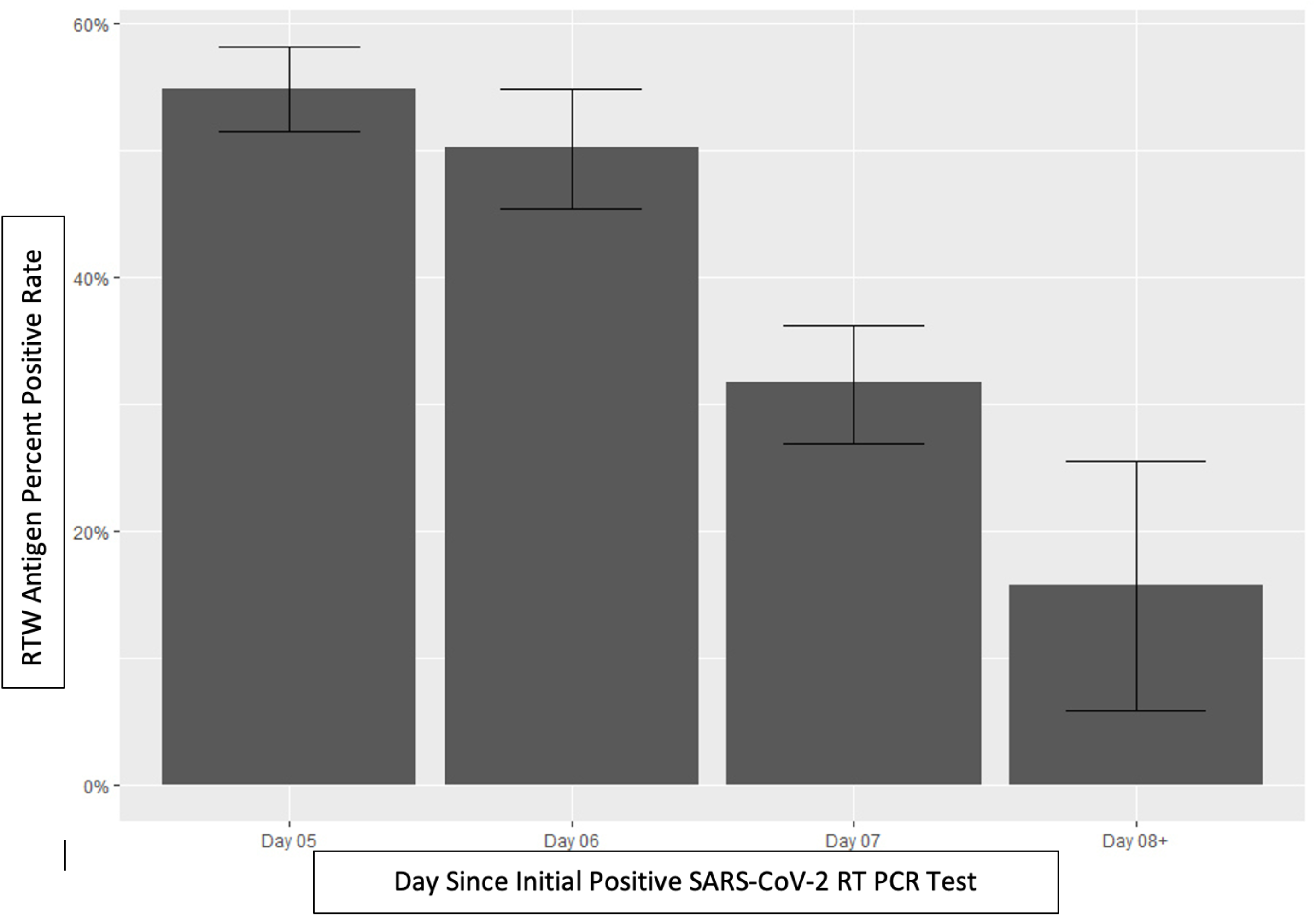
RTW antigen test percent positivity rate with 95% confidence interval by number of days since initial SARS-CoV-2 RT PCR test.

Each demographic characteristic analyzed by the chi-squared test was included in the logistic regression model after demonstrating a significant association with antigen results. In the multivariate model, a positive RTW antigen result was more likely in vaccinated (OR: 1.41, 95%CI: 1.00-1.98, p <0.05), boosted for more than 90 days (OR: 2.21, 95%CI: 1.56-3.12, p<0.0001), and boosted within 90 days (OR: 2.08, 95% CI: 1.34-3.24, p < 0.01) compared to not being vaccinated. A positive RTW antigen result was less likely on day 7 (OR: 0.39, 95%CI: 0.29-0.51, p<0.0001) and after at least 8 days (OR: 0.16, 95%CI: 0.08-0.35, p<0.0001) compared to day 5. Neither gender nor age were significantly associated with the antigen test result in the multivariate model after adjusting for covariates.

## Discussion

We found that a high percentage of HCWs remained positive on their day 5 and 6 RTW antigen test, consistent with findings by other groups ^6,9–11^. With growing data suggesting that a positive COVID-19 antigen test correlates closely with shedding of culturable SARS-CoV-2 virus and likely with contagiousness, these findings merit a re-assessment of the guidance on the optimal time to return HCWs back to work after COVID-19 infection, especially if a RTW antigen strategy is not employed ^12–15^. The use of RTW antigen testing in our setting likely prevented infectious employees from returning to work, thereby reducing the risk of continued transmission within the healthcare setting that further depletes the workforce and could contribute to nosocomial transmission to patients.

A significant decline in the odds of a positive RTW antigen was noted from day 7 and beyond following a positive PCR test. In settings where RTW antigen tests are not readily available due to testing capacity or supply shortages, extending the duration of HCW work restriction to at least 7 days during staffing shortages may reduce the number of possibly contagious HCWs returning to work and thus limit transmission events that would contribute to further staffing loss and exposure of vulnerable patients.

While our study was conducted in the context of returning HCWs to work, these results may have implications for community guidance as well. Current CDC guidance for the community outlines a 5-day isolation period following symptom onset or a positive test if symptoms are improving, followed by masking when around others for an additional 5 days ^9^. In the community setting, where masking is often inconsistent, modifying guidance to extend the initial isolation period from 5 to 7 days after a positive test may decrease overall community transmission ^16,17^.

In the community study from the Yukon-Kuskokwim Delta region, persons who were unvaccinated and without prior infection were more likely to have a positive follow-up COVID-19 antigen test when compared to vaccinated or previously infected individuals ^6^. An unexpected finding from our study was that vaccinated and boosted HCWs had significantly higher odds of testing positive on their RTW antigen test— independent of the timing of the RTW test and accounting for other demographic variables— when compared to their unvaccinated colleagues (Table 2). This unusual signal was also identified by Tande et al who reported that HCWs who were up to date on their COVID-19 vaccinations were more likely to be positive on their first RTW antigen test compared to those who were unvaccinated or not up-to-date on their vaccines ^18^. The study by Landon and colleagues further noted that boosted HCWs were more likely to have a positive RTW antigen test than their non-boosted peers ^5^. To our knowledge, ours is the largest to date in HCWs for which other important potential confounding factors are controlled for.

**TABLE 2:** Multivariable logistic regression of factors associated with a positive RTW antigen test.

|  | Odds Ratio (95% CI) | p-value | Standard Error |
| --- | --- | --- | --- |
| **Gender (ref = Male)** |  |  |  |
| Female | 0.84 (0.66-1.06) | 0.15 | 0.12 |
| **Age group, yrs (ref = 18-29)** |  |  |  |
| 30-39 | 0.92 (0.70-1.21) | 0.54 | 0.14 |
| 40-49 | 1.21 (0.90-1.65) | 0.21 | 0.16 |
| 50+ | 1.31 (0.99-1.74) | 0.06 | 0.14 |
| **Vaccination Status (ref = Not Vaccinated)** |  |  |  |
| Vaccinated* | 1.41 (1.00-1.98) | < 0.05 * | 0.17 |
| Boosted 90+ Days | 2.21 (1.56-3.12) | <0.0001*** | 0.18 |
| Boosted <90 Days | 2.08 (1.34-3.24) | < 0.01 ** | 0.23 |
| **Days from Initial PCR Test (ref = Day 5)** |  |  |  |
| Day 6 | 0.85 (0.68-1.07) | 0.16 | 0.12 |
| Day 7 | 0.39 (0.29-0.51) | <0.0001 *** | 0.14 |
| Days 8+ | 0.16 (0.08-0.35) | <0.0001 *** | 0.40 |

\*completion of primary series

The reasons for our finding of a higher proportion of positive RTW antigen in our vaccinated and boosted HCWs are unclear and may reflect a yet unaccounted-for characteristic(s) of that population including differences in prior infection between the groups. There is ample data demonstrating that being up-to-date on COVID-19 vaccination is protective against severe disease and is associated with decreased risks of hospitalization, the need for intensive care or mechanical ventilation, and death from COVID-19 ^19–23^. Thus, this unexpected finding needs to be investigated further by leveraging larger data repositories that may have information on comorbidities, prior COVID infections and vaccine status with the need for results being carefully messaged to avoid undermining ongoing and future vaccination efforts.

## Limitations

This observational study was conducted in a single health system in southwest VA when the omicron variant and its subvariants were predominant and thus may not be generalizable to other settings or applicable to other variants. As with most retrospective studies we recognize that our findings could be due to unobserved confounders. However, other studies done during the omicron surge in different healthcare settings seem to signal a similar conclusion.

We did not conduct any viral cultures to determine virus viability in the antigen positive HCWs and thus cannot draw definite conclusions about their infectiousness. Nonetheless, recently published studies have found a good correlation between positive antigen tests, low PCR cycle thresholds ^24^ and culturable virus^13,14^ which are the more established measures of infectiousness, making our findings worth further evaluation.

## Conclusion

Our findings suggest that a high proportion of HCWs have a positive RTW antigen for at least 6 days following their initial positive SARS-CoV-2 PCR test and are could still be contagious, even when symptoms are improving. During contingency staffing, guidance for returning HCWs to work should include antigen testing where feasible or delaying return until at least 7 days from the initial positive PCR test. This could support more robust staffing levels by minimizing transmission within the healthcare setting. COVID-19 isolation guidance for the community should also be reviewed and extended if these findings are replicated especially using follow-up viral cultures. The unexpected finding that the vaccinated and boosted HCW had higher odds of a positive RTW antigen test compared to unvaccinated HCW also merits further investigation.

## Data Availability

All data produced in the present study are available upon reasonable request to the authors

## Notes

### Competing Interest Statement

The authors have declared no competing interest.

### Funding Statement

This study did not receive any funding

### Author Declarations

This study was deemed exempt by the Carilion Clinic IRB.

## References

1. Centers for Disease Control and Prevention. Interim Guidance for Managing Healthcare Personnel with SARS-CoV-2 Infection or Exposure to SARS-CoV-2. Published 2022. https://www.cdc.gov/coronavirus/2019-ncov/hcp/guidance-risk-assesment-hcp.html

2. Centers for Disease Control and Prevention. Strategies to Mitigate Healthcare Personnel Staffing Shortages. Published 2022. https://www.cdc.gov/coronavirus/2019-ncov/hcp/mitigating-staff-shortages.html

3. Pekosz A, Parvu V, Li M, et al. Antigen-Based Testing but Not Real-Time Polymerase Chain Reaction Correlates With Severe Acute Respiratory Syndrome Coronavirus 2 Viral Culture. Clinical Infectious Diseases. 2021;73(9):e2861–e2866. doi:10.1093/cid/ciaa1706

4. Lopera TJ, Alzate-Ángel JC, Díaz FJ, Rugeles MT, Aguilar-Jiménez W. The Usefulness of Antigen Testing in Predicting Contagiousness in COVID-19. Microbiol Spectr. 2022;10(2):e01962–21. doi:doi:10.1128/spectrum.01962-21

5. Landon E, Bartlett AH, Marrs R, Guenette C, Weber SG, Mina MJ. High Rates of Rapid Antigen Test Positivity After 5 days of Isolation for COVID-19. medRxiv. Published online January 1, 2022:2022.02.01.22269931. doi:10.1101/2022.02.01.22269931

6. Lefferts B, Blake I, Bruden D, et al. Antigen Test Positivity After COVID-19 Isolation - Yukon-Kuskokwim Delta Region, Alaska, January-February 2022. MMWR Morb Mortal Wkly Rep. 2022;71(8):293–298. doi:10.15585/mmwr.mm7108a3

7. Chu VT, Schwartz NG, Donnelly MAP, et al. Comparison of Home Antigen Testing With RT-PCR and Viral Culture During the Course of SARS-CoV-2 Infection. JAMA Intern Med. 2022;182(7):701–709. doi:10.1001/jamainternmed.2022.1827

8. Korenkov M, Poopalasingam N, Madler M, et al. Evaluation of a Rapid Antigen Test To Detect SARS-CoV-2 Infection and Identify Potentially Infectious Individuals. J Clin Microbiol. 2021;59(9):e0089621. doi:10.1128/JCM.00896-21

9. Boucau J, Marino C, Regan J, et al. Duration of Shedding of Culturable Virus in SARS-CoV-2 Omicron (BA.1) Infection. New England Journal of Medicine. 2022;387(3):275–277. doi:10.1056/NEJMc2202092

10. Landon E, Bartlett AH, Marrs R, Guenette C, Weber SG, Mina MJ. High Rates of Rapid Antigen Test Positivity After 5 days of Isolation for COVID-19. medRxiv. Published online 2022:2022.02.01.22269931. doi:10.1101/2022.02.01.22269931

11. Tande AJ, Swift MD, Challener DW, et al. Utility of Follow-up Coronavirus Disease 2019 (COVID-19) Antigen Tests After Acute Severe Acute Respiratory Syndrome Coronavirus 2 (SARS-CoV-2) Infection Among Healthcare Personnel. Clinical Infectious Diseases. Published online 2022. doi:10.1093/cid/ciac235

12. Pekosz A, Parvu V, Li M, et al. Antigen-Based Testing but Not Real-Time Polymerase Chain Reaction Correlates With Severe Acute Respiratory Syndrome Coronavirus 2 Viral Culture. Clin Infect Dis. 2021;73(9):e2861–e2866. doi:10.1093/cid/ciaa1706

13. Lopera TJ, Alzate-Ángel JC, Díaz FJ, Rugeles MT, Aguilar-Jiménez W. The Usefulness of Antigen Testing in Predicting Contagiousness in COVID-19. Microbiol Spectr. 2022;10(2):e0196221. doi:10.1128/spectrum.01962-21

14. Chu VT, Schwartz NG, Donnelly MAP, et al. Comparison of Home Antigen Testing With RT-PCR and Viral Culture During the Course of SARS-CoV-2 Infection. JAMA Intern Med. 2022;182(7):701–709. doi:10.1001/jamainternmed.2022.1827

15. Boucau J, Marino C, Regan J, et al. Duration of Shedding of Culturable Virus in SARS-CoV-2 Omicron (BA.1) Infection. N Engl J Med. 2022;387(3):275–277. doi:10.1056/NEJMc2202092

16. Centers for Disease Control and Prevention. Isolation and Precautions for People with COVID-19. Published 2022. https://www.cdc.gov/coronavirus/2019-ncov/your-health/isolation.html?CDC_AA_refVal=https%3A%2F%2Fwww.cdc.gov%2Fcoronavirus%2F2019-ncov%2Fyour-health%2Fquarantine-isolation.html

17. Fischer CB, Adrien N, Silguero JJ, Hopper JJ, Chowdhury AI, Werler MM. Mask adherence and rate of COVID-19 across the United States. PLoS One. 2021;16(4):e0249891. doi:10.1371/journal.pone.0249891

18. Tande AJ, Swift MD, Challener DW, et al. Utility of Follow-up COVID-19 Antigen Tests After Acute SARS-CoV-2 Infection Among Healthcare Personnel. Clin Infect Dis. Published online March 30, 2022. doi:10.1093/cid/ciac235

19. Sheikh A, McMenamin J, Taylor B, Robertson C. SARS-CoV-2 Delta VOC in Scotland: demographics, risk of hospital admission, and vaccine effectiveness. The Lancet. 2021;397(10293):2461–2462. doi:10.1016/S0140-6736(21)01358-1

20. Arbel R, Hammerman A, Sergienko R, et al. BNT162b2 Vaccine Booster and Mortality Due to Covid-19. N Engl J Med. 2021;385(26):2413–2420. doi:10.1056/NEJMoa2115624

21. Altarawneh HN, Chemaitelly H, Ayoub HH, et al. Effects of Previous Infection and Vaccination on Symptomatic Omicron Infections. N Engl J Med. 2022;387(1):21–34. doi:10.1056/NEJMoa2203965

22. Watson OJ, Barnsley G, Toor J, Hogan AB, Winskill P, Ghani AC. Global impact of the first year of COVID-19 vaccination: a mathematical modelling study. Lancet Infect Dis. Published online June 23, 2022. doi:10.1016/S1473-3099(22)00320-6

23. Center for Disease Control and Prevention. Benefits of Getting A COVID-19 Vaccine. Published 2022. https://www.cdc.gov/coronavirus/2019-ncov/vaccines/vaccine-benefits.html

24. Routsias JG, Mavrouli M, Tsoplou P, Dioikitopoulou K, Tsakris A. Diagnostic performance of rapid antigen tests (RATs) for SARS-CoV-2 and their efficacy in monitoring the infectiousness of COVID-19 patients. Sci Rep. 2021;11(1):22863. doi:10.1038/s41598-021-02197-z

