## Appendix A for "When should healthcare workers with COVID-19 return to work? An analysis of follow-up antigen test results after a positive COVID test"

### Employee Return-to-Work Timeline following a Positive COVID-19 Test

- This timeline is applicable to employees who completed a self-attestation, received a red boarding pass, and were tested for COVID-19 at a Carilion Testing Center.
- Employees are responsible for following the steps below and updating their manager as they move through the process.

Day 0 is the date of initial positive PCR or antigen test collection

| Days 0-4 | Days 5-7 | Days 8-10 | Day 11 |
|---|---|---|---|
| Remain out of work through Day 4 → | On Days 5-7 self-evaluate symptoms:<br>Have you been free from diarrhea and vomiting for 48 hours AND fever free for 24 hours without the use of fever-reducing medication, AND other symptoms (e.g. cough) are improving? | | |
| | **If Yes,** proceed to a Carilion testing site for return-to-work antigen testing. Orders will automatically be available on Days 5-7. Employees who test negative will be cleared to return to work by the testing center. → | Employees who test positive on their return-to-work antigen test will remain out of work through Day 10. No additional testing will be done. → | Return to work on Day 11 |
| | **If No,** criteria for return-to-work antigen testing is not met. Remain out of work. → | Remain out of work through Day 10. On Day 10 self-evaluate symptom:<br>Have you been free from diarrhea and vomiting for 48 hours AND fever free for 24 hours without the use of fever-reducing medication, AND other symptoms (e.g. cough) are improving? → | **If Yes,** return to work on Day 11<br><br>**If, No** follow routine attendance processes with your manager to return to work. Contact your primary care provider for prolonged duration of symptoms. |

Note: There may be instances where employees may not follow this timeline. They will be addressed on a case-by-case basis.

Updated 1/12/22
